# Identification of Novel mRNA Biomarkers with Improved Performance for Colorectal Cancer Screening from a Multicenter Large Gene Screen

**DOI:** 10.64898/2026.02.03.26345497

**Authors:** Loren Hansen, Houcong Liu, Haijiu Lin, Changpu Song, Yanyang Liang, Jakob Kirchner, Dan Chen, Zhufang Chen, Jihui Du, Wenying Pan

**Affiliations:** El Capitan Biosciences, 7068 Koll Center Pkwy, Suite 402, Pleasanton, CA 94566; Research Center for Clinical and Translational Medicine, Shenzhen Nanshan People’s Hospital and Affiliated Nanshan Hospital of Shenzhen University, Shenzhen, Guangdong 518052, China; Guangdong Jiyin Biotech, D3 Building, TCL international E city, no. 1001, Zhong Shan Yuan Road, Nanshan District. Shenzhen, China

## Abstract

**Background:** Colorectal cancer (CRC) is a leading cause of cancer mortality. While early detection improves outcomes, current non-invasive tests often lack sensitivity for early-stage CRC and advanced precancerous lesions (APL). Stool-based host messenger RNA (mRNA) biomarkers offer a promising approach, though the most clinically useful candidates remain undefined.

**Methods:** We screened for mRNA biomarkers by first using bioinformatic analysis of tissue RNA-seq datasets to identify candidate genes with strong and ubiquitous differential expression in CRC versus normal tissues. The top 135 computationally predicted biomarkers were evaluated using "gold standard" RT-PCR on clinical stool samples across two independent cohorts.

**Results:** Several biomarkers, including *PPBP*, *MYC*, *MMP7*, and *TGFBI*, exhibited strong predictive power. Integrating top-performing markers through machine learning yielded an AUC of 0.98 for CRC and 0.76 for APL detection. The optimized panel demonstrated 98% sensitivity for CRC and 50% for APL, with a specificity of 90%.

**Conclusions:** This study derives a high-performance mRNA-based stool test for non-invasive CRC screening. Our findings demonstrate that a multi-marker panel achieves exceptional sensitivity and good specificity, providing a viable tool for clinical diagnostics.

## Background

Globally, Colorectal cancer (CRC) is the third most commonly diagnosed cancer and the second most common cause of cancer-related deaths for both men and women. CRC is therefore a serious global health burden ^1^. Colorectal cancer almost always follows the adenoma-carcinoma pathway in which a series of stepwise mutations and epigenetic modifications slowly transform normal epithelium to dysplastic epithelium; this transformation commonly takes 10-15 years ^2^. The slow progression from benign polyps to malign growths provides an opportunity to identify and remove advanced precursor lesions (APL) or early stage malignant tumors. Given the slow development from benign polyps to malign growths, CRC is an attractive target for prevention and early detection through population-based screening programs. Indeed colorectal cancer screening is one of the few cancers in which screening has been shown to lead to a significant reduction of incidence rates and mortality ^3^ with compliance with screening being one of the key drivers of screening effectiveness ^4^. Colonoscopies are the gold standard approach to screen for precursor lesions and early stage disease with excellent sensitivity and specificity ^5^ screening every ten years with a colonoscopy has been shown to have the largest reduction in mortality of the various screen options ^3^. Unfortunately compliance with a colonoscopy is frequently not optimal due to the invasive nature of the procedure, the unpleasant preparation, and time requirements ^6^. In addition, colonoscopy resources in developing countries are frequently not sufficient to support population level CRC screening ^7^.

To address these issues, non-invasive screening is an increasingly common alternative. The most common non-invasive test worldwide is detection of occult blood in stool ^2,5,8^. Fecal immunochemical tests (FIT) tests are cost effective, have high sensitivity and specificity in detecting the presence of blood proteins in stool and do not require dietary restrictions. However, FIT, while it has good specificity for CRC/APL detection at ∼90-95%, suffers from poor sensitivity in APL detection (10-40%) and has modest sensitivity in detection of early stage CRC tumors (37-70%) ^5,9–11^.

Alternate strategies that depend on the exfoliation of tumor cells into the colon-rectal lumen have been developed. The most mature approach is a multitarget test pairing DNA methylation with somatic mutations and FIT ^12^. Detection of tumor/APL-derived mRNA from exfoliated cells, instead of DNA, has a number of potential advantages. Aberrantly expressed mRNA may have many hundreds or thousands of copies per shed tumor cell, potentially increasing the sensitivity of detection, especially for small lesions. There have been a number of studies demonstrating that measuring CRC associated mRNA in stool can be a reliable method to detect colorectal cancers and advanced adenomas. ^13,14^. Indeed, on the basis of a large-scale clinical validation trial in the intended use population, the FDA recently approved a mRNA-based test for CRC/APL population-based screening ^15^.

However, which mRNA biomarkers are best suited for CRC/APL screening has yet to be identified or validated. In previous work ^16^ we showed that by taking advantage of the thousands of high quality, publicly available RNA-seq tissue transcriptomics datasets, clinically useful stool-based mRNA biomarkers can be predicted. Here we extend that work and perform the largest screen to date using individual PCR of stool-based mRNA biomarkers (Fig 1).

**Figure 1:**
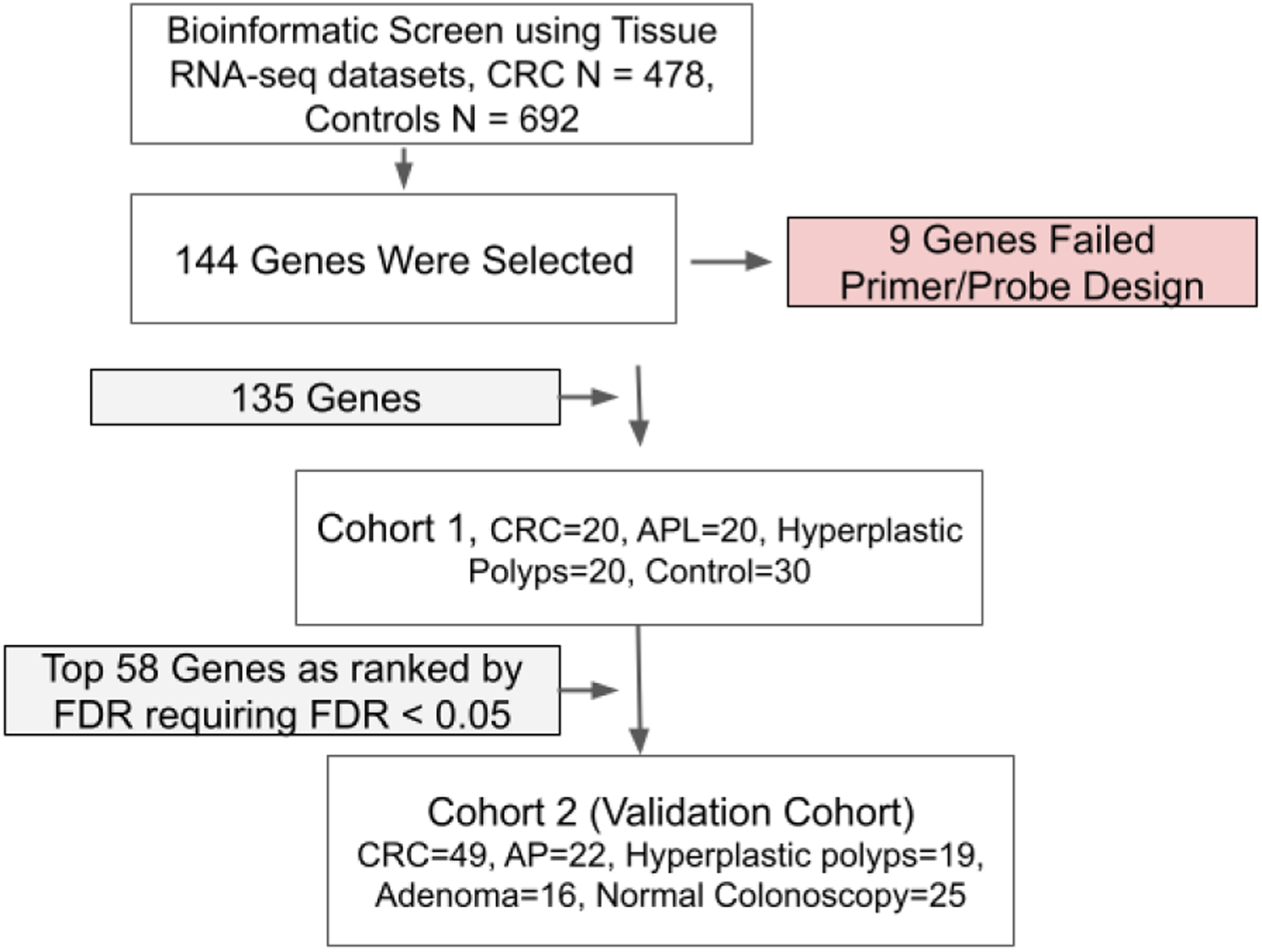
Study Flowchart.

## Materials and Methods

### Tissue Biomarker Screen

The majority of genes tested (129) were obtained from the bioinformatic screen as described in our previous manuscript16. Briefly, CRC transcriptomic datasets and paired normal tissue samples were downloaded from The Cancer Genome Atlas (TCGA) database 19 and healthy colon controls were downloaded from the Genotype-Tissue Expression (GTEx) database. The combined TCGA/GTE dataset consisted of 478 colon cancer tissue samples and 692 normal colon/rectum tissue samples. The data consisted of gene count matrices.

Batch correction on the downloaded data to merge the healthy controls from the TCGA database with the GTEx normal samples was performed using combat-seq20. A differential expression analysis comparing gene expression in CRC tissue vs healthy colon tissue was then performed using edgeR21. In addition to the differential expression analysis a variety of metrics was calculated for each gene. These metrics included the percentile ranking of each gene’s median expression level in CRC and healthy tissue, and each gene’s area under the curve (AUC) comparing gene expression in CRC tissue to healthy tissue controls. The differential expression false discovery rate (FDR) and log fold change were calculated using edgeR. The AUC and median percentile ranks in normal/disease tissue was calculated using custom code written in R.

Stool is a complex matrix with microbial RNA being the dominant source of RNA in stool. It is expected that RNA derived from cancer cells will commonly constitute a small fraction of the total RNA present in stool. Therefore, additional filters were applied to identify and rank genes in terms of their ability to detect the presence of abnormal cells in stool. Based on comparisons between CRC and normal tissue samples, we hypothesized that genes showing strong and consistent differential expression across most tumors relative to healthy controls would be strong candidates for stool-based biomarkers in CRC and APL screening. In addition, genes that are strongly expressed in CRC tissue are likely easier to detect in stool when the fraction of pathogenic cells is small. The following methodology was used to obtain these genes: differential expression FDR<0.001 and AUC > 0.9 and a log base 2-fold change difference in expression between CRC and healthy tissue > 2. The genes that passed this filter were then sorted by median expression in CRC samples from highest to lowest. The top of this list would be genes ubiquitously differentially over-expressed across many CRC tissue samples compared to normal tissue samples and that are also highly expressed in CRC tissue.

Including the GTEx samples enabled us to greatly expand the pool of normal colon/rectal tissue samples. We also ran the bioinformatic analysis using only TCGA samples to avoid any potential batch effects. The large majority of genes overlapped between the two analyses but there were 15 that were unique to the TCGA only analysis. We also included these genes in the set of genes to be screened on clinical samples. The resulting gene list was 144 genes.

### Experimental Screen Design

To test the clinical performance of the computationally predicted biomarkers and to narrow down the set of biomarkers further, a screen was performed first testing the computationally predicted biomarkers on cohort 1 (90 clinical samples) then testing the best performing genes from cohort 1 on an independently collected sample cohort. TaqMan assays for each tested gene were designed in-house to be RNA specific by placing either one of the primers or the probe across a splice junction. For 9 of the genes, suitable primers/probes could not be designed and these genes did not move forward to be tested on clinical samples. The 135 genes with suitable TaqMan assays were then tested on cohort 1 which consisted of 90 samples (CRC N=20, APL N=20, hyperplastic polyps (HP) N=20 and control samples N=30). Control samples for cohort 1 consisted of volunteers without a colonoscopy. Given the low incidence rate of colorectal cancer the control samples for cohort 1 are unlikely to have undiagnosed colorectal cancer, but a number of advanced adenomas and adenomas are likely present.

The genes from cohort 1 were ranked based on their diagnostic performance. The Mann-Wilcox test was performed for each gene comparing the normalized gene expression values of the CRC patients compared to the controls (control + hyperplastic polyp samples). The calculated p-values were multiple testing corrected using false discovery rates (FDR) and genes were ranked according to their FDR. The top ranked 58 genes were then tested on the independently collected validation cohort 2. Sample cohort 2 was collected through a commercial biobank in China and confirmed colonoscopy negative samples were obtained for this cohort. Cohort 2 consists of the following sample breakdown: total number of samples N=131, CRC N=49, APL N=22, hyperplastic polyps N=19, adenoma N=16, normal colonoscopy N=25.

### Data Presentation and Statistical Analysis

The delta-delta Ct method 22 was used to calculate the relative gene expression. Normalized gene expression data was used for all analysis. The delta-delta Ct method, commonly written as 2^–ΔΔCt, enables comparison of relative fold change in gene expression between samples by normalization to a reference gene.

First, the delta Ct is calculated by subtracting for each gene the reference gene’s Ct value, i.e., ΔCt = Ct_target – Ct_reference. The ΔCt of the reference samples are then subtracted from the ΔCt of each sample, i.e., ΔΔCt = ΔCt_sample – ΔCt_reference_samples. The fold change comparing each sample to the reference sample is then calculated as fold_change = 2^(–ΔΔCt). The delta-delta Ct fold change represents differences in gene expression normalized by a reference gene and compared to a set of reference samples. Commonly the reference samples used in delta-delta Ct normalization would be the control samples. But for some of our genes, none of the control samples had detectable Ct values to be used as references. So instead the reference samples used in the delta-delta Ct calculation was the mean Ct value for those cancer samples with detectable signal defined as Ct values < 40 cycles. The reference gene used was GAPDH. Hence for our analysis, delta-delta Ct is gene expression relative to the mean gene expression observed in the cancer samples.

All machine learning experiments (Tables 3 and 4 and Figures 3 and 4) were performed using the tidymodels package in R and default parameters were used for the random forest model. Classification performance was estimated using an average of five-fold cross validations repeated 30 times with random splits of the data for each cross validation experiment. Five-fold cross validation consists of randomly splitting the data into 5 roughly equal groups, training the model on the combination of four of the groups and testing the model on the last remaining group. This process is repeated until all groups have been used as the held out test set. The input data to the machine learning experiments was the normalized Ct values. For those genes with undetectable transcripts in a particular stool sample the normalized Ct value was set to 0.

The control group for cohort 2 samples was the combination of normal colonoscopy stool samples, stool samples from patients with hyperplastic polyps and stool samples from patients with non-advanced adenomas. The control group for cohort 1 samples was stool collected from volunteers and stool from patients with hyperplastic polyps. Sensitivity and specificity was calculated using a 0.5 class probability cutoff. If the reported class probability was > 0.5 then the sample was predicted to be a disease sample, and if the class probability was < 0.5 then the sample was predicted to be a control sample.

To adjust the specificity of the mRNA panel to match the specificity observed for FIT (Table 3 last row) a more rigorous class probability cutoff of 0.57 was used. For this analysis, if the reported class probability was > 0.57 then the sample was predicted to be a disease sample, and if the class probability was < 0.57 then the sample was predicted to be a control sample.

Statistical significance testing of differential representation of each of the genes (Supplemental tables 1 and 3) comparing the CRC or APL patient samples with control samples was performed using the Mann-Whitney test, p-values were adjusted for multiple testing comparisons using the Benjamini Hochberg method, (False Discovery Rate) FDR <0.05 was used as the significant cutoff.

### Gene Panel Selection

The panel of genes was selected by using cohort 1 samples to select the optimal gene panel; this gene panel was then tested on cohort 2 samples. A greedy algorithm was used to identify the gene panel. Genes were ranked by APL AUC based on their performance in cohort 1. Genes were added one at a time to the gene panel based on their ranking. Each gene combination was tested on cohort 1 samples using the random forest machine learning algorithm and five-fold cross validation. The final gene panel was selected based on the point when adding more genes did not substantially change the performance. (Fig 3).

### Patients and Sample Collection

#### Cohort 1 Sample Collection

Cohort 1 samples were collected in Shenzhen Nanshan People’s Hospital. Sample collection occurs ≥14 days prior to colonoscopy or ≥14 days after diagnostic colonoscopy (prior to therapeutic/surgical intervention). Colonoscopy findings meeting one of the following three categories-Colorectal Cancer (CRC): Newly diagnosed colorectal cancer, stages I-IV; Advanced Adenoma (APL):

Meeting one of the following criteria: Tubular or serrated adenomatous polyp ≥10 mm or ≥ 5 tubular or serrated adenomas. Villous adenoma, or tubulovillous adenoma with >25% villous component. Adenoma with high-grade intraepithelial neoplasia (HGIN); Hyperplastic Polyp(s) (HP): Subjects with <20 hyperplastic polyps.

Control samples consisted of volunteers who were recruited from the pool of patients undergoing health examinations at Shenzhen Nanshan People’s Hospital during the same time period colorectal samples were being collected.

Volunteers did not undergo colonoscopies, but the following criteria was used to reduce the risk of including patients with colorectal lesions in the control group: Normal fecal characteristics, negative fecal occult blood tests, and normal blood tests and tumor marker assessments.

Fresh morning fecal samples (10–15 g) were collected using sampling rods, placed into fecal collection tubes, and sealed. Samples were aliquoted within 2 hours and stored at −80°C for future analysis.

The study of cohort 1 was approved by the Ethics Committee of Shenzhen Nanshan People’s Hospital (Approval No. KY-2020-045-01). Informed consent was given before stool was collected. The research was performed in accordance with the relevant guidelines and regulations, including the Declaration of Helsinki.

#### Cohort 2 Sample Collection

Collection of human clinical stool samples was done through a CRO (Contract Research Organization) in China which used an observational method to obtain human stool samples and associated clinical data from symptomatic and asymptomatic patients. Patient samples were collected either prospectively or retrospectively. Clinical samples were sent to Tellgen (Shanghai, China) for FIT testing.

Fresh stool samples (15–40 g) were collected by patients, and sealed into fecal collection tubes. Samples were sent to the hospital within 4 hours and stored at −20°C. All subjects were aged 43 years or older and scheduled for colonoscopy. Sample collection occurs ≥14 days prior to colonoscopy or ≥14 days after diagnostic colonoscopy (prior to therapeutic/surgical intervention).

Colonoscopy findings meeting one of the following five categories-Colorectal Cancer (CRC): Newly diagnosed colorectal cancer, stages I-IV; Advanced Precancerous Lesions (APL): Meeting one of the following criteria: Tubular or serrated adenomatous polyp ≥10 mm. ≥ 5 tubular or serrated adenomas. Villous adenoma, or tubulovillous adenoma with >25% villous component. Adenoma with high-grade intraepithelial neoplasia (HGIN); Healthy Control (HC) were subjects with normal colonoscopy findings; Hyperplastic Polyp(s) (HP): Subjects with <20 hyperplastic polyps; Adenomatous Polyp(s) (Adenoma): Meeting one of the following criteria: Tubular adenoma(s) <10 mm in diameter. Serrated adenomatous polyp(s) <10 mm. ≥20 hyperplastic polyps.

Samples were excluded for the following reasons: presence of specific diseases or conditions including inflammatory bowel disease (IBD), Celiac disease without dietary control, acute diarrhea, or gastrointestinal infections other than intestinal tuberculosis; Any other malignancy of the digestive system, history of any malignancy diagnosed within the past 5 years; Hereditary colorectal cancer syndromes (e.g., Hereditary Nonpolyposis Colorectal Cancer (HNPCC/Lynch Syndrome), Familial Adenomatous Polyposis (FAP)); History of colorectal resection for any reason other than sigmoid diverticula or bowel resection >1 meter. For non-CRC control groups, samples were excluded for the following reasons: Lack of complete colonoscopy examination results or female subjects who are pregnant or breastfeeding.

Upon receipt, stool samples were aliquoted and stored under two conditions: one half aliquoted and directly frozen at −80°C, and the other half aliquoted and stored in Zymo Research DNA/RNA Shield™ preservation buffer before freezing at –80°C.

This study of cohort 2 was approved by the following Ethics Committees: Ethics committee/IRB of Jiangmen Central hospital (Approval No.江心医伦理审查 【2024】228号A), YiXing People’s Hospital (Approval No. 伦审2024科031-01). Informed consent was given before stool was collected. The research was performed in accordance with the relevant guidelines and regulations, including the Declaration of Helsinki.

### RNA Isolation and RT-PCR

RNA was extracted from 0.25 g of stool using the Omega Bio-Tek E.Z.N.A.® Stool RNA Kit (R6828), which employs a phenol-chloroform extraction method. To remove potential PCR inhibitors, the Zymo Research OneStep™ PCR Inhibitor Removal Kit (D6035) was used following the RNA extraction. The extracted RNA was resuspended in 50 µL of DEPC-treated water per tube. RNA concentration and purity were measured using a NanoDrop spectrophotometer. The typical RNA yield was approximately 450 µg per sample. The RNA was then aliquoted and stored at −80°C until further use.

For RT-qPCR, Takara One Step PrimeScript™ III RT-qPCR Mix with UNG (RR601A) was utilized for RT-qPCR. Each reaction was set up with a total volume of 10 µL, including 2 µL of extracted RNA. The reactions were performed on an ABI 7500 Real-Time PCR System with the following thermal cycling conditions: reverse transcription at 50°C for 5 minutes, initial denaturation at 95°C for 10 seconds, followed by 40 cycles of 95°C for 10 seconds and 60°C for 34 seconds.

The stool RNA isolation and RT-qPCR were performed by partnering with a local laboratory in China (Fapon biotech, Shenzhen, China).

## Results

### Cohort 1 Results

We performed a bioinformatic screen to identify the top genes that, based on their expression in tissue (CRC N= 478, Healthy Controls N= 692), were most promising as stool-based CRC screening biomarkers (See Materials and Methods). The top 135 computationally predicted genes were then tested on cohort 1 clinical samples (Figure 1, Material and Methods).

A set of performance metrics was calculated for each gene (Supplemental excel table 1, Material and Methods). In total there were 4 genes with an AUC >= 0.9 for CRC detection (OLR1, TGFBI, CXCL8, and TIMP1). The number of genes with a statistically significant (False Discovery Rate (FDR) < 0.05) difference in level of expression comparing CRC with control samples was 77 out of 135 genes, which translates to a ∼57% successful prediction rate for the predictions based on tissue expression data. The top performing gene for APL detection was MMP7 with an AUC of 0.74. MMP7 also has a CRC AUC of 0.83 and represents a well-balanced biomarker with good CRC and APL diagnostic performance.

**Table 1:** Performance across cohorts of top 5 genes as ranked by CRC AUC on cohort 1 samples.

| **Gene Name** | **CRC AUC Cohort 1** | **CRC AUC Cohort 2** | **APL AUC Cohort 1** | **APL AUC Cohort 2** |
| --- | --- | --- | --- | --- |
| OLR1 | 0.93 | 0.95 | 0.58 | 0.66 |
| TGFBI | 0.92 | 0.93 | 0.68 | 0.67 |
| CXCL8 | 0.91 | 0.97 | 0.60 | 0.74 |
| TIMP1 | 0.90 | 0.94 | 0.64 | 0.72 |
| PPBP | 0.88 | 0.97 | 0.67 | 0.69 |

We then investigated if there were biological processes in common among the top performing genes in cohort 1. Genes were ranked according to their FDR and those genes with a FDR < 0.05 (N=77) underwent a biological process enrichment analysis using the Panther gene list enrichment analysis tool ^17^, apoptotic regulation was found to be the most commonly enriched biological process in the cohort 1 top performing genes (Supplemental excel table Go_Biological_process_analysis).

### Cohort 2 Results

To determine the reproducibility of the clinical performance, 58 of the top performing genes based on cohort 1 were measured on 131 independently collected clinical stool samples (Figure 1).

A set of performance metrics was calculated for each gene and is given in supplemental table 3. The pattern of differential expression observed in colorectal cancer samples compared to controls was highly reproducible across clinical cohorts. Of the 58 genes tested with statistically significant differential expression comparing CRC stool samples to control stool samples in cohort 1, 57 out of the 58 genes had statistically significant differential expression in the cohort 2 samples (defined as a FDR < 0.05) (See supplemental table 3).

The “big picture” was highly reproducible with most genes in both cohorts having statistically significant differential expression comparing CRC stool to stool from control patients. The exact clinical performance observed was more variable with a number of genes having significant differences in clinical performance comparing cohort 1 to cohort 2 (Figure 2 panels A and B).

**Figure 2:**
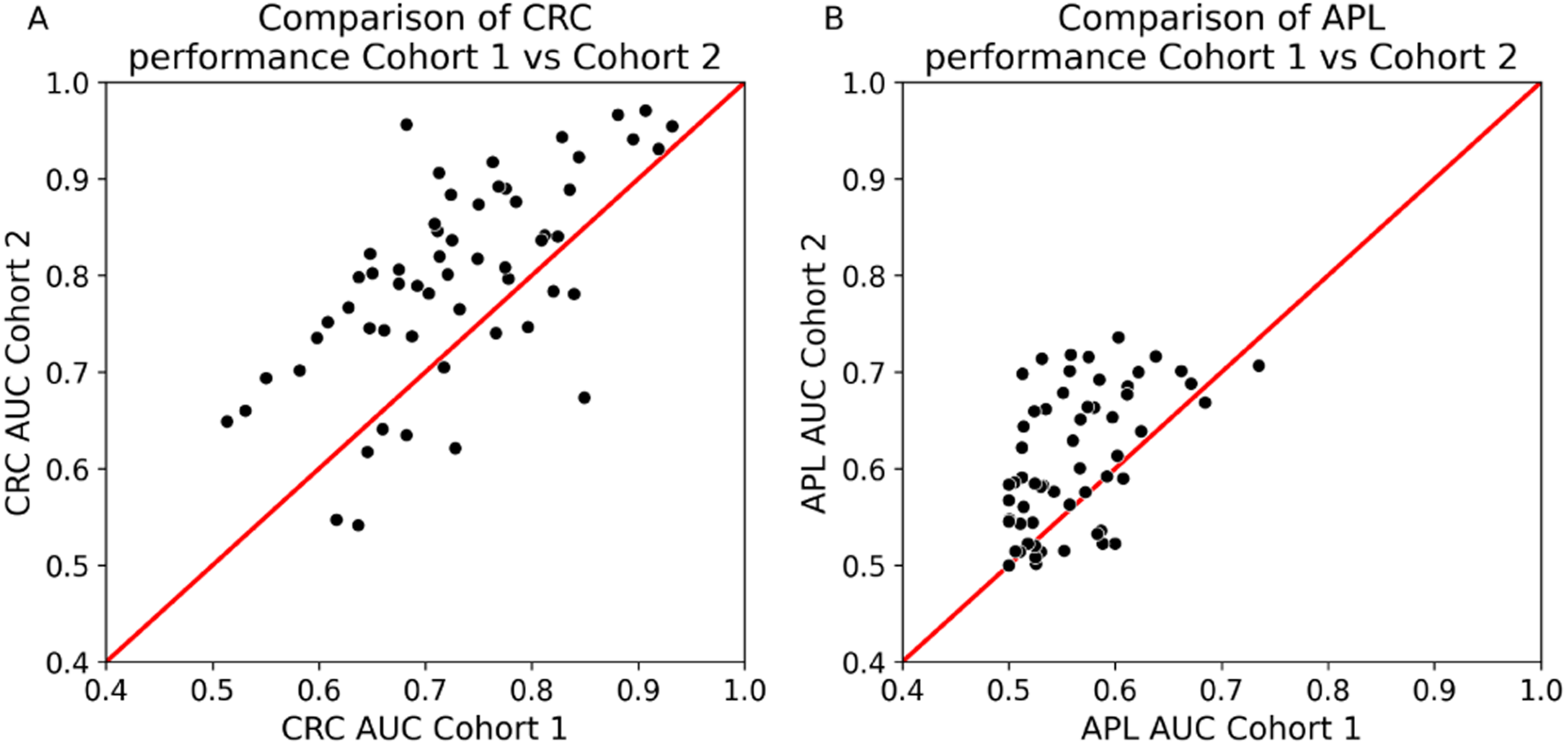
Comparison of CRC clinical performance between cohort 1 and cohort 2 samples. **Panel A:** Each dot represents a gene. The y-axis is the CRC AUC measured in cohort 2 samples and the x-axis is the CRC AUC measured in cohort 1 samples. Every dot above the diagonal represents a gene whose CRC AUC was higher in cohort 2 compared to cohort 1. Those dots below the diagonal represent genes whose CRC AUC was lower in cohort 2 samples compared to cohort 1. **Panel B:** Comparison of APL clinical performance between cohort 1 and cohort 2 samples. The y-axis is the APL AUC measured in cohort 2 samples and the x-axis is the APL AUC measured in cohort 1 samples.

The correlation in CRC AUC between cohort 1 and cohort 2 was moderate with a correlation coefficient of 0.69 (Pearson coefficient, p-value = 2.1 e-9). While many genes did have significant variance in clinical performance comparing the two cohorts, the top performing genes had less variance with similar good performance across the sample cohorts (Table 1).

There was a correlation between CRC and APL performance with a correlation coefficient of 0.54 (Pearson coefficient, p-value = 1.04 e-5) where those genes performing well at APL identification also have a tendency to perform well at CRC identification (Supplemental Figure 1). Interestingly, gene expression across samples was overall highly correlated (supplemental figure 2)

The reproducibility in APL performance was lower than the reproducibility in CRC performance with a correlation coefficient comparing cohort 1 and cohort 2 APL AUCs of 0.48 (Pearson correlation, p-value = 9.9 e-5). Similar to CRC performance, the top performing APL genes had higher consistency in performance between the two sample cohorts (supplemental table 4).

### Gene Combinations

To test how well combinations of genes would perform we selected a panel of genes to test. Genes in cohort 1 were ranked by their APL AUC and then added one at a time to test the performance of each gene combination on cohort 1. The final gene panel was selected by adding genes until the performance stops improving (Figure 3). Performance for each gene combination was obtained using the random forests machine learning algorithm ^18^ and 5-fold cross validation (see Materials and Methods).

The CRC performance was relatively stable with a sharp increase in performance combining the top 4 genes. Adding additional genes shows little improvement in performance (Figure 3 CRC curve). The APL performance, however, was more variable. This is likely because, in general, the APL signal was a good deal weaker than the CRC signal, with higher variance in performance between sample cohorts for the top performing APL genes (Figure 3 APL curve).

**Figure 3:**
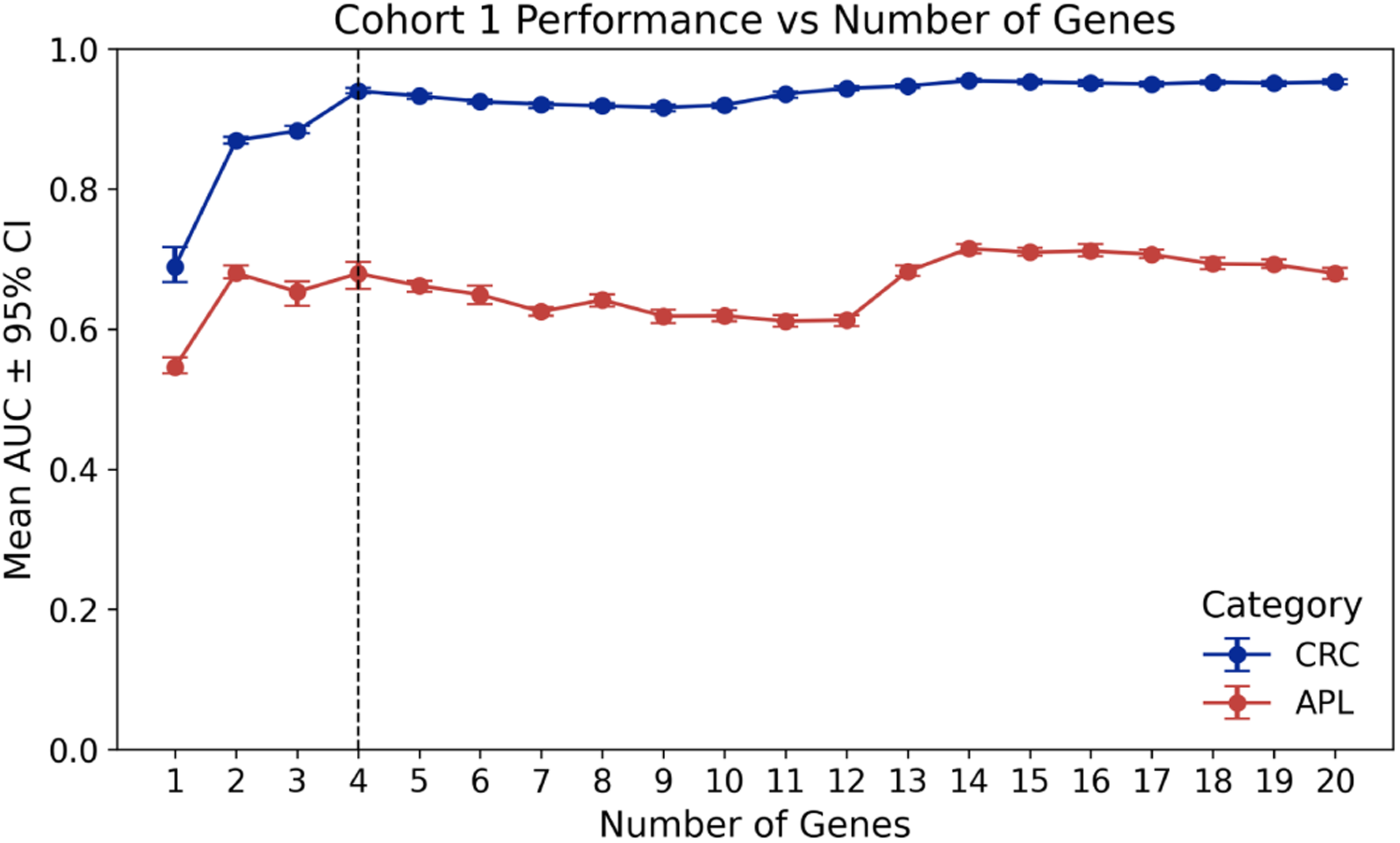
Plotted is the AUC for CRC and APL as calculated on cohort 1 for different gene combinations. Genes were ranked by APL performance on cohort 1, then genes were added one at a time based on their ranking and the performance was calculated for each gene combination using 5-fold cross validation and a random forest model. This process was repeated 20 times; each point represents the mean AUC across all 20 iterations. The x-axis is the number of genes in the gene combination being tested. The y-axis is the performance of each gene combination. The vertical line represents the cutoff used to select the gene panel.

Building a panel for use in the clinic is a balance between cost, ease of use and performance. The smaller the gene panel, the better. Examining the plots of CRC/APL performance versus number of combined genes, a 4-gene panel looks like the best combination of performance versus gene number after taking into account both APL and CRC diagnostic performance. The top 4 genes had very consistent performance across both cohort 1 and cohort 2 (table 1 and Figure 4 panels A and B)

The top 4 genes were combined and tested on the independently collected cohort 2 (Genes used: PPBP, MYC, MMP7 and TGFBI). The AUC using the combined 4 genes tested on cohort 2 samples was: 0.98 CRC AUC and 0.76 APL AUC, the sensitivity for CRC detection was 98%, the APL sensitivity was 50% and the specificity was 90%. The performance of the gene panel tested on cohort 1 samples was: 0.94 CRC AUC and 0.67 APL AUC the sensitivity for CRC detection was 85% the APL sensitivity was 45% the specificity was 92% (see Table 2, and Fig 4). Importantly, performance did not greatly differ based on stage and early stage lesions were as likely to be detected as later stages (see Supplemental Table 3).

**Table 2:** Performance across cohorts of mRNA gene panel and FIT.

| Biomarkers Tested | CRC Sensitivity | APL Sensitivity | Specificity |
| --- | --- | --- | --- |
| Cohort 1- mRNA Panel | 85% | 45% | 94% |
| Cohort 2 - mRNA Panel | 98% | 50% | 90% |
| Cohort 2 - FIT | 86% | 13.6% | 93% |
| Cohort 2- mRNA<br>(Specificity set to 93%) | 98% | 50% | 93% |

**Figure 4.**
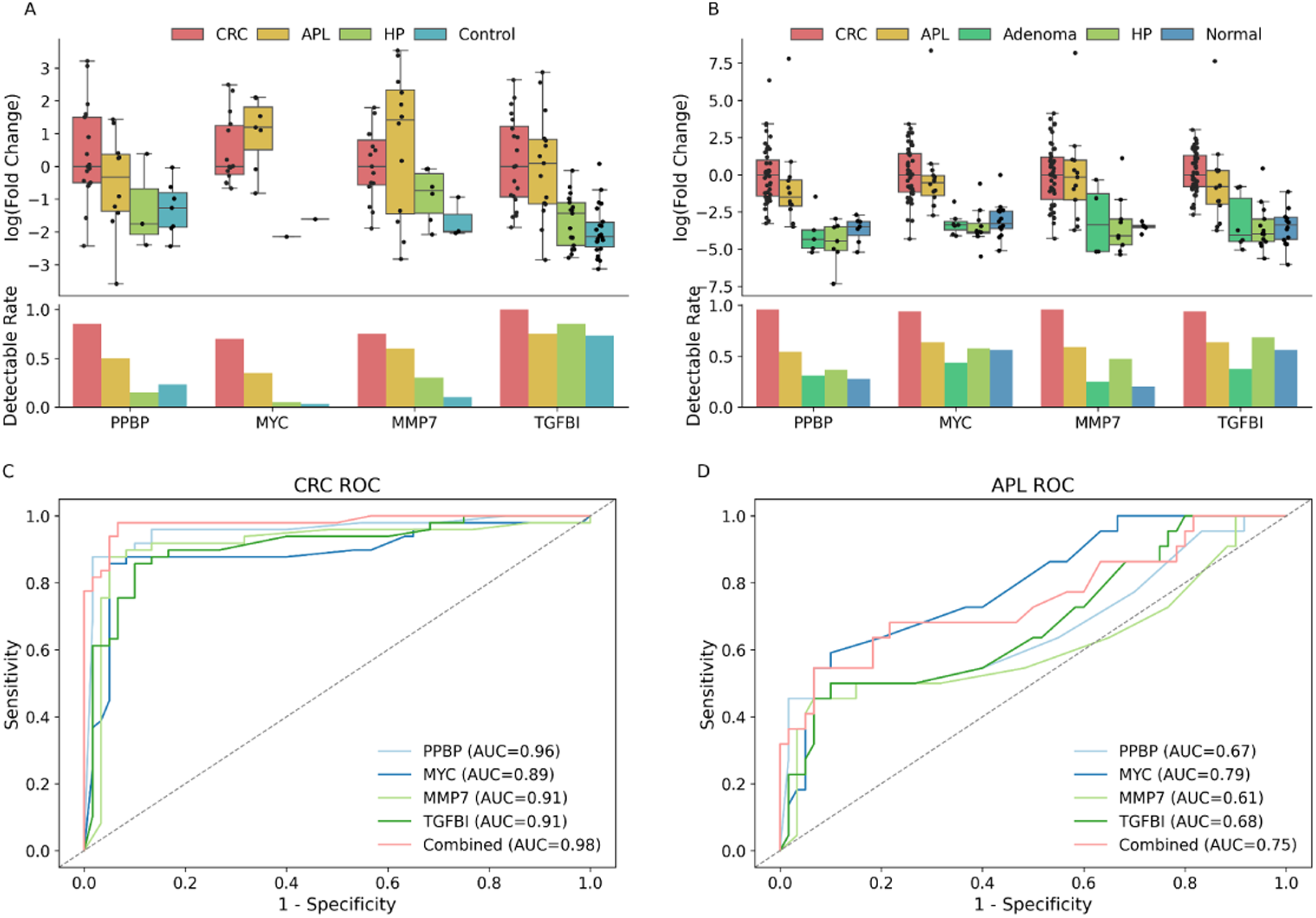
**Panel A:** Plotted delta-delta Ct normalized values in cohort 1 stool samples for the 4 gene mRNA panel. Every dot represents a sample with detectable transcripts; the top plot is the normalized Ct values represented by fold change. The bar plot represents the fraction of samples with detectable copy numbers in stool. Only those samples with detectable Ct values are plotted in the top panel. **Panel B:** Plotted delta-delta Ct normalized values in cohort 2 stool samples for the gene mRNA biomarker panel. Every dot represents a sample with detectable transcripts; the top plot is the normalized Ct values represented by fold change. The bar plot represents the fraction of samples with detectable copy number in stool. Only those samples with detectable Ct values are plotted in the top panel. **Panel C:** ROC plot of CRC versus controls, each gene is a different color. The combined performance is represented by the solid black line. **Panel D:** ROC plot of APL versus controls, each gene is a different color. The combined performance is represented by the solid black line.

As a point of comparison, FIT was performed for all samples in cohort 2. FIT had a CRC sensitivity of detection of 86%, a sensitivity of APL detection of 13.6% and a specificity of 93%. To fairly compare performance of the different methods, the specificity of the mRNA panel was adjusted to match the specificity of FIT. This was done by adjusting the class probability cutoff, such that 93% of the control samples (HP + Normal colonoscopy + Non-advanced adenomas) would be correctly classified as controls (see Materials and Methods). At a specificity of 93% the mRNA panel’s performance did not change, i.e. the sensitivity of CRC detection remained 98%, the sensitivity of detection for APLs was also unchanged at 50% (See Table 2).

## Discussion

A systematic and comprehensive investigation of mRNA biomarkers for the purposes of CRC screening has not yet been done. In a previous study ^16^ we tested the top 20 bioinformatically predicted genes with good results. In this study we build on our earlier work by greatly extending the number of genes tested on clinical samples from 20 to 135 genes and adding another independently collected sample cohort. In general, the bioinformatic predictions based on tissue expression were moderately accurate with a ∼57% success rate.

Our results indicate a high degree of reproducibility in clinical performance across clinical cohorts for the best performing CRC genes with the reproducibly decreasing for genes with moderate performance. Advanced adenomas had a lower degree of reproducibility; this is likely due to the overall lower signal for APL samples compared to CRC samples and may also reflect differences in APL sample composition (size, type and location of lesions) between sample cohorts. Location and type of APLs can have a strong impact on ease of detection ^23^. The trend for the best performing genes to have higher reproducibility across sample cohorts also held true for APL performance.

In general gene expression as measured in stool was strongly correlated across samples (supplemental figure 2). A higher degree of cellular shedding will increase the detectable expression level for all host-derived genes. The strong correlation observed across stool samples suggests the degree of cell shedding may be a more important determinant of measured expression in stool than the level of expression within the cells being shed.

In the 4 gene panel derived from our analysis (MYC, MMP7, PPBP, TGFBI), two out of the 4 (MYC, MMP7), in agreement with our results, have been shown in previous studies to be differentially overexpressed in the stool of CRC patients relative to controls ^24–27^.

Our top performing genes therefore represent a mix of genes that have previously been shown in the literature to be differentially expressed in the stool of CRC patients and novel genes discovered by our biomarker screen. MYC is a well-known and well-studied driver of cancer genesis and growth and its misregulation is a key factor of CRC initiation and progression ^28^. MYC protein plays multiple roles in processes ranging from cell growth, proliferation and immune evasion ^28^.

MMP7 is a secreted endopeptidase that has been linked to many different cancers, including CRC ^29^. When expressed by cancer cells MMP7 protein may inhibit apoptosis ^30^ and cell adhesion ^29^. However, MMP7 is also commonly overexpressed in inflamed tissue ^31^. So MMP7 expression may be a combination of expression in tumor cells and in immune cells.

TGFBI encodes a secreted extracellular matrix protein involved in numerous processes, including cell adhesion, migration, wound healing and tissue remodeling ^32,33^. Similar to MMP7, TGFBI plays an important role in regulating the immune response (de Streel and Lucas, 2021; Massagué and Sheppard, 2023) and is expressed in several different immune cell types ^36^.

PPBP encodes a chemokine that plays a significant role in immune cell recruitment and is also involved in tissue regeneration and angiogenesis ^37^. It is overexpressed in several cancers and also in chronic inflammatory conditions, such as IBD and rheumatoid arthritis ^37^.

The genes in our gene panel are multifunctional and play many roles, but 3 out of the 4 are strongly linked with immune response (MMP7, TGFBI and PPBP). Defense response was also one of the enriched GO terms that the best performing genes in cohort 1 had in common (supplemental table 2). This suggests that the immune system’s interaction with malignant tissue may be one of the strongest sources of diagnostic signal. The immune signature in our sample cohorts appears to be relatively specific to patients with neoplastic or advanced precancerous lesions. But further testing on patients known to be suffering from colitis is necessary.

Limitations of this study include biased sample collection (enriched for rectal cancers), relatively small clinical sample size, older samples (in some cases ∼2 years old), and that the samples collected were not the intended use screening population. In particular the control group for cohort 1 were volunteers without colonoscopy negative confirmation. Given the low incidence rate of colorectal cancer in the general population, it is unlikely the control group included a patient with undiagnosed colorectal cancer. But given the prevalence of APLs, it is likely that undiagnosed APLs were present in the control group. Undiagnosed precancerous lesions may have caused some genes to appear less specific than they truly are since any samples with a high level of signal in the control group were treated as a false positive in the analysis.

## Conclusion

In general the results of our large biomarker screen are quite promising. The best performing genes have reproducible clinical performance across independently collected sample cohorts. Combining the top performing genes in cohort 1 and testing them on an independent dataset we get clinical performance that is competitive with the most advanced stool-based tests currently available ^23^. Out of the CRC samples collected, the 4 gene panel detected 48 out of the 49 colorectal tumors at a respectable 90% specificity. When directly compared with FIT at the same specificity, the 4 gene mRNA panel had 12% higher sensitivity for CRC detection and 37% higher sensitivity for APL detection.

All current multi-target nucleic acid based panels used for CRC screening pair FIT with a panel of nucleic acid-based biomarkers. Combining a protein assay with a nucleic acid panel can complicate the patient’s experience in collecting stool samples and increases the difficulty of automating the workflow. Our panel of 4 genes does not require FIT and consists of a relatively straightforward 4 gene multiplex assay similar to multiplex assays currently in widespread use for infectious disease detection using stool.

## Data Availability

Processed results are contained in supplemental excel files. Raw data may be
available upon request.

## Additional Information

### Authors’ contributions

*Concept and Clinical Study Design-* Jihui Du, Houcong Liu, Dan Chen, Haijiu Lin

*Patient recruitment and sample collection (Cohort 1):* Houcong Liu, Zhufang Chen, Xiaohong Zhang, Jihui Du

*Biomarker discovery: Loren Hansen, Wenying Pan*

*Data Analysis software development*: Loren Hansen, Wenying Pan

*Clinical Data Analysis*: Yanyang Liang

*Drafting of the Manuscript*: Loren Hansen, Houcong Liu

*Wet lab method development and processing samples*: Changpu Song, Haijiu Lin, Dan Chen and Jakob Kirchner.

### Ethics Approval

This study was approved by the Ethics Committees of all hospitals at which samples were collected. Informed consent was given before stool was collected. The research was performed in accordance with the relevant guidelines and regulations, including the Declaration of Helsinki.

### Consent for Publication

All authors have approved the manuscript for submission. And the content of this manuscript has not been published elsewhere.

### Availability of data and materials

Processed results are contained in supplemental excel files. Raw data may be available upon request.

## Acknowledgement

We appreciate Pinjie Lei’s contribution in code review and reviewing the manuscript.

