## supplemental document for "Identification of Novel mRNA Biomarkers with Improved Performance for Colorectal Cancer Screening from a Multicenter Large Gene Screen"

**Supplementary Information**

**Supplemental Table 2:** GO Enrichment Analysis of biological results

| **GO biological process complete** | **Fold Enrichment** | **FDR** |
| --- | --- | --- |
| positive regulation of mesenchymal cell apoptotic process (GO:2001055) | > 100 | 4.41E-02 |
| cellular response to UV-A (GO:0071492) | 70.16 | 4.51E-02 |
| mesenchymal cell apoptotic process (GO:0097152) | 64.31 | 4.49E-02 |
| regulation of mesenchymal cell apoptotic process (GO:2001053) | 59.37 | 3.33E-02 |
| response to UV-A (GO:0070141) | 51.45 | 4.10E-02 |
| extracellular matrix disassembly (GO:0022617) | 22.37 | 3.77E-02 |
| bone morphogenesis (GO:0060349) | 13.12 | 4.56E-02 |
| skeletal system morphogenesis (GO:0048705) | 7.93 | 3.91E-02 |
| cellular response to lipid (GO:0071396) | 5.24 | 5.54E-02 |
| defense response to symbiont (GO:0140546) | 3.7 | 4.47E-02 |
| positive regulation of cellular component organization (GO:0051130) | 3.47 | 3.99E-02 |
| cellular response to chemical stimulus (GO:0070887) | 2.76 | 3.82E-02 |
| animal organ development (GO:0048513) | 2.3 | 4.12E-02 |
| positive regulation of cellular process (GO:0048522) | 1.81 | 3.82E-02 |
| positive regulation of biological process (GO:0048518) | 1.81 | 1.04E-01 |

**Table 2:** Genes were ranked by CRC vs control differential expression FDR in cohort 1: Genes with a FDR < 0.05 were selected for a GO enrichment analysis (N=76).

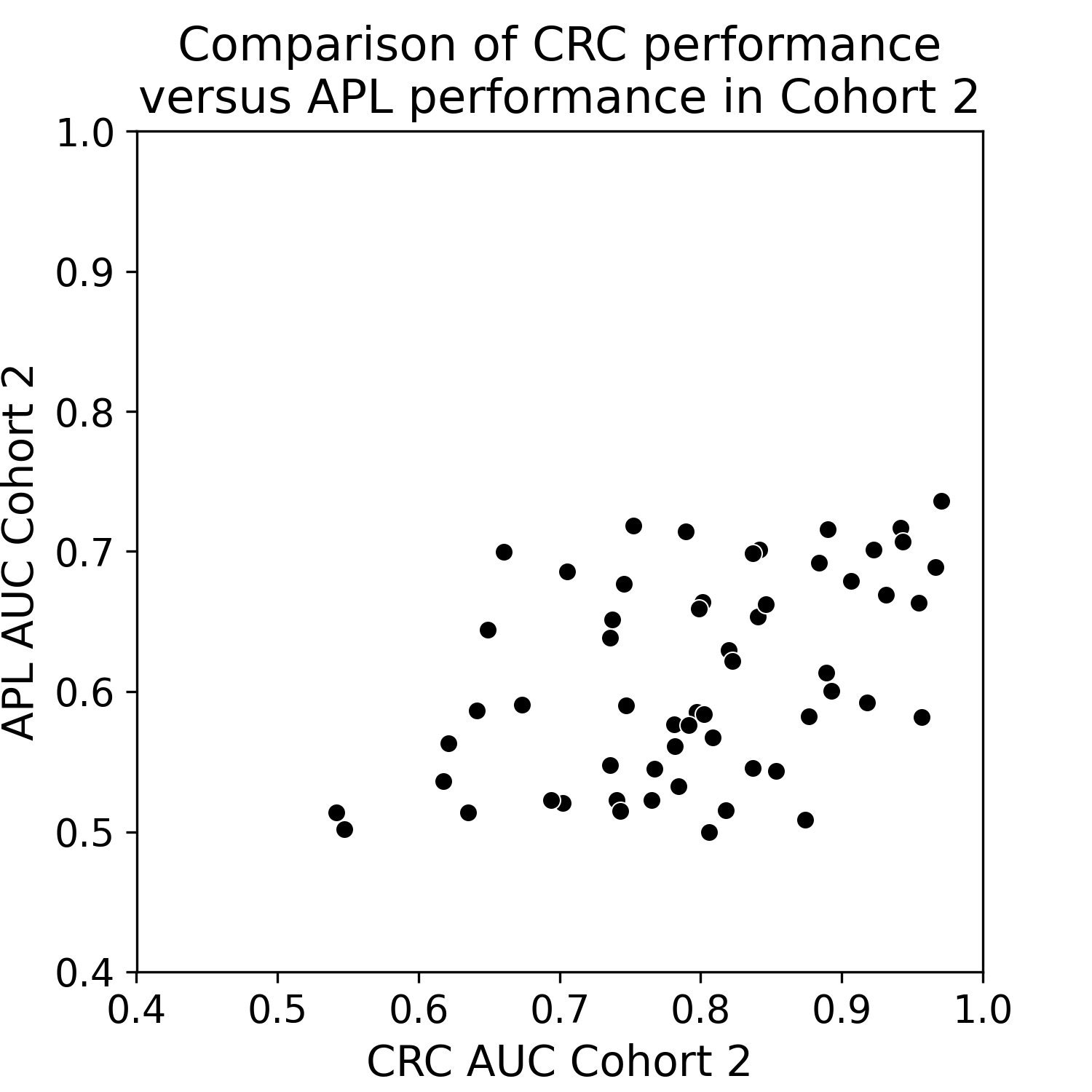

**Supplemental Figure 1: Comparison of CRC vs APL performance in cohort 2 samples.** Each dot represents a gene. The y axis is the APL AUC measured in cohort 2 the x axis is the CRC AUC measured in cohort 2. There is a correlation between CRC and APL performance.

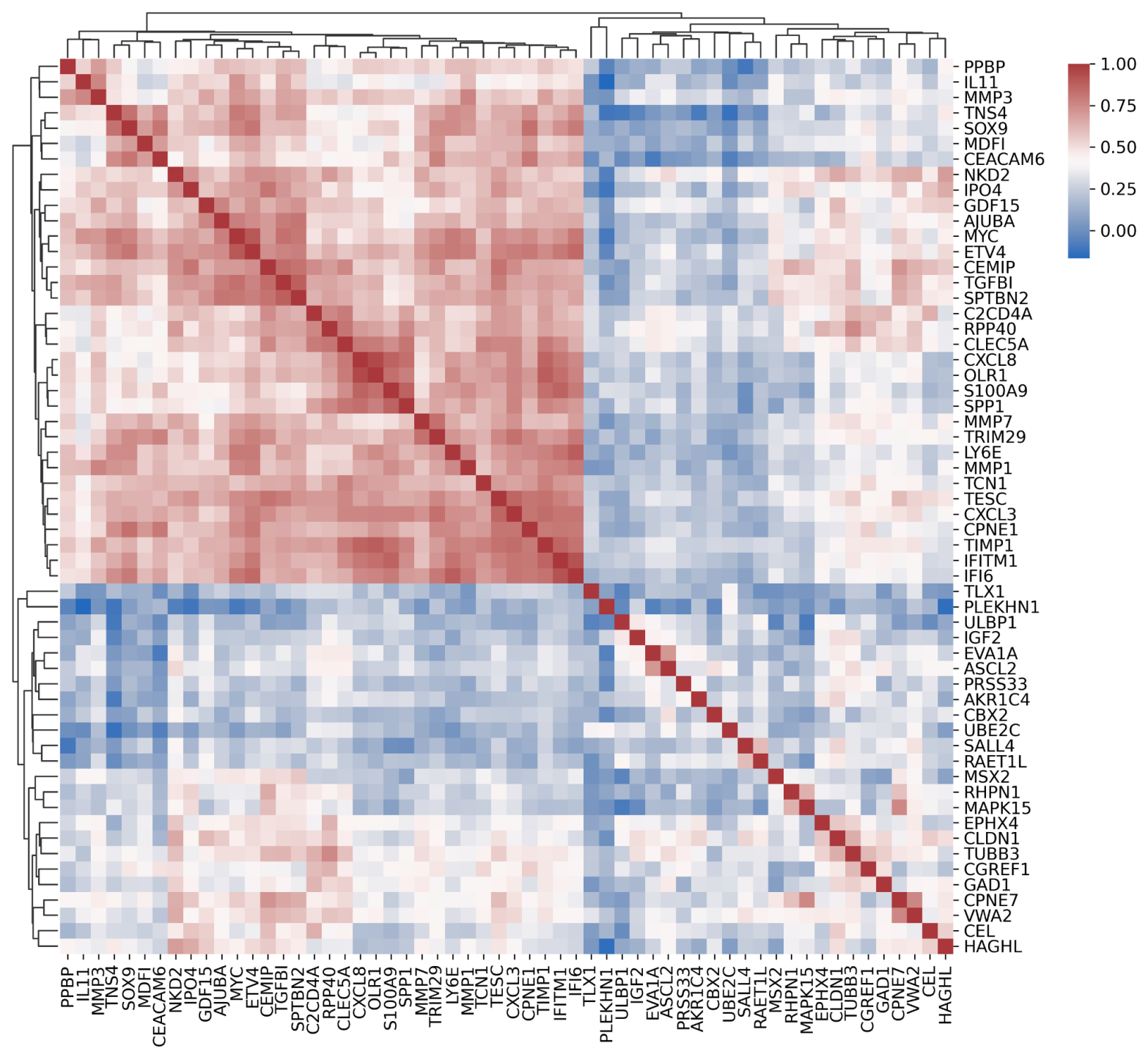

**Supplemental Figure 2: Correlation Matrix.**

Each cell in the matrix represents the correlation between two genes. Correlation between each gene expression was calculated for cohort 2 CRC samples using spearman. In general most genes show a significant correlation. Those genes with little correlation with other genes are in general genes with no detectable expression in most samples (See supplemental table 3).

| **Gene  Name** | **CRC  AUC Cohort 1** | **CRC  AUC Cohort 2** | **APL  AUC Cohort 1** | **APL  AUC Cohort 2** |
| --- | --- | --- | --- | --- |
| MMP7 | 0.83 | 0.94 | 0.74 | 0.71 |
| TGFBI | 0.92 | 0.93 | 0.68 | 0.67 |
| PPBP | 0.88 | 0.97 | 0.67 | 0.69 |
| MYC | 0.84 | 0.92 | 0.66 | 0.70 |
| TIMP1 | 0.90 | 0.94 | 0.64 | 0.72 |

**Supplemental Table 4: Performance across cohorts of top 5 genes as ranked by APL AUC on cohort 1 samples.**

| **Stage** | **Number of Samples** | **Sensitivity** | **Specificity** |
| --- | --- | --- | --- |
| I | 7 | 100% | 90% |
| II | 16 | 100% | 90% |
| III | 20 | 100% | 90% |
| IV | 5 | 80% | 90% |

**Supplemental Table 5: Performance of mRNA gene panel in cohort 2 categorized by stage.**
